# Persons with multiple sclerosis reveal distinct kynurenine pathway metabolite patterns: a multinational cross-sectional study

**DOI:** 10.1101/2025.02.20.25322520

**Authors:** M. Kupjetz, M. Langeskov-Christensen, M. Riemenschneider, S. Inerle, U. Ligges, T. Gaemelke, N. Patt, J. Bansi, R. Gonzenbach, M. Reuter, F. Rosenberger, T. Meyer, A. McCann, P.M. Ueland, S.F. Eskildsen, M.K.E. Nygaard, N. Joisten, L.G. Hvid, U. Dalgas, P. Zimmer

## Abstract

Neuroaxonal damage in multiple sclerosis (MS) results from an interplay of neurotoxic pathomechanisms combined with a reduced neuroprotective capacity of neurons and glia to resist neurotoxic damage. Kynurenine pathway (KP) imbalance resembles some of the molecular mechanisms central to the incompletely understood MS pathophysiology. To study the role of the KP in MS, we performed targeted metabolomics on serum samples from 353 persons with MS and 111 healthy individuals, and detected MS-specific differences in concentrations of most kynurenines. Using exploratory factor analysis, we then identified two distinct KP metabolite patterns: the inflammation-driven neurotoxic pattern „NeuroTox“ and the neuroprotective pattern „NeuroPro“. Our results show that greater „NeuroTox“ and lower „NeuroPro“ were associated with higher disease severity. The novelty of our data-driven approach that identified distinct KP metabolite patterns in MS advocates for future studies using comparable approaches to investigate whether KP imbalance follows similar disease-specific patterns in diseases other than MS.

## Introduction

Multiple sclerosis (MS) is a chronic inflammatory and neurodegenerative disease of the central nervous system (CNS) characterized by focal inflammatory lesions and diffuse, smoldering disease activity. The resulting neuroaxonal damage is thought to emerge from an interplay of multiple neurotoxic pathomechanisms, combined with a reduced neuroprotective capacity of neurons and glia to resist neurotoxic damage. Understanding the complex interactions between molecular mechanisms and the various neuroimmunological pathways that contribute to the neurotoxic CNS microenvironment in MS has recently been highlighted as key to increasing neuronal resilience and slowing disease progression.^1^

The kynurenine pathway (KP) of tryptophan (Trp) metabolism (**Figure 1A**) may represent one of the neuroimmunological pathways involved in the neurotoxic CNS microenvironment associated with the pathophysiology of MS.^2^ Trp metabolites, collectively termed kynurenines, do not only serve as precursors in nicotinamide adenine dinucleotide (NAD^+^) *de novo* synthesis but also modulate inflammatory activity, neuronal and glial homeostasis, and neurotransmission. Moreover, the consequences of KP imbalance partly resemble the molecular mechanisms currently discussed as central to MS pathophysiology (e.g., glutamate excitotoxicity).^3^

**Figure 1.**
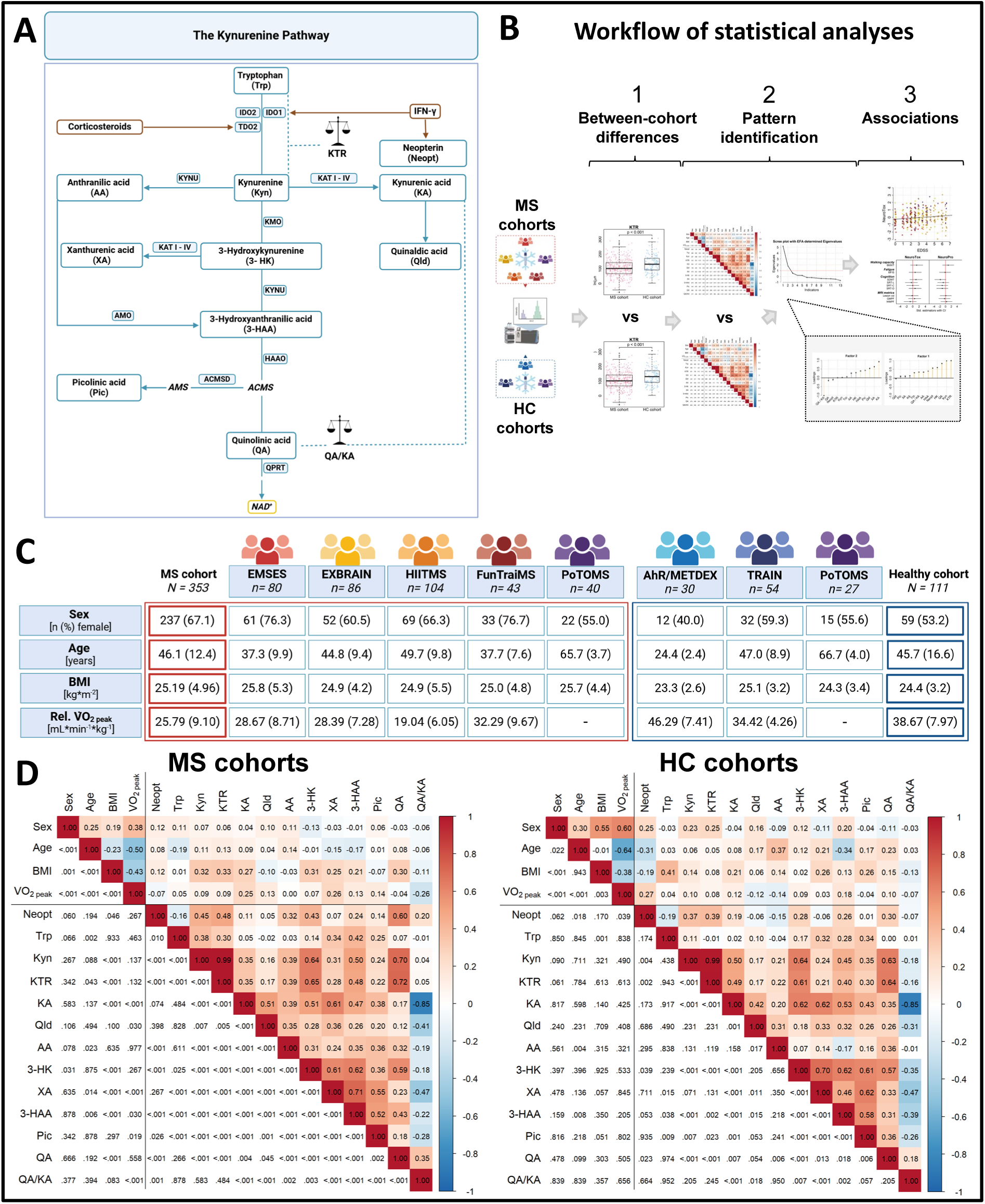
Study overview and results of correlation analysis. **A** Schematic illustration of the kynurenine pathway. Neopterin and the kynurenine-to-tryptophan ratio indicate inflammatory stimulation of the kynurenine pathway, as interferon-gamma induces both the activation of the enzyme indoleamine 2,3-dioxygenase and the formation of neopterin by activated macrophages and dendritic cells. **B** Workflow of statistical analyses. **C** Characteristics of the pooled multiple sclerosis and healthy cohort with individual subcohorts. Categorical data are given as total number and percentage (%). Continuous data are given as mean (SD). Cardiorespiratory fitness (relative V̇O_2 peak_) was not assessed in the PoTOMS study. V̇O_2 peak_ was missing for individual participants of the FunTraiMS (n = 4), EXBRAIN (n = 2), TRAIN (n = 2), and AhR/METDEX (n = 1) studies. **D** Heatmaps showing correlations between participant characteristics and kynurenine pathway metabolites/ratios, presented separately for the pooled multiple sclerosis cohort (left) and healthy cohort (right). Pearson’s correlation analysis was partialized for sex, age, body mass index, and relative V̇O_2 peak_. The lower triangles of the heatmaps show false discovery rate-adjusted p-values. The upper triangles of the heatmaps show Pearson’s r correlation coefficients. AA = anthranilic acid; BMI = body mass index; HC = healthy cohort; KA = kynurenic acid; KTR = kynurenine-to-tryptophan ratio; Kyn = kynurenine; MS = multiple sclerosis; Neopt = neopterin; Pic = picolinic acid; Trp = tryptophan; QA = quinolinic acid; QA/KA ratio = quinolinic acid-to-kynurenic acid ratio; Qld = quinaldic acid; V̇O_2 peak_ = peak oxygen consumption during cardiopulmonary exercise testing divided by kilograms body weight; XA = xanthurenic acid; 3-HAA = 3-hydroxyanthranilic acid; 3-HK = 3-hydroxykynurenine.

According to the traditional view, KP imbalance is thought to result from chronic inflammatory stimulation of the KP, as indicated by an elevated kynurenine-to-tryptophan ratio (KTR), together with a shift of KP activity toward a neurotoxic branch, primarily characterized by the accumulation of the neurotoxic KP metabolite quinolinic acid (QA). QA exerts its neurotoxic effects via several molecular mechanisms, such as N-methyl-D-aspartate receptor (NMDAR)-mediated glutamate excitotoxicity or the promotion of oxidative stress.^4^ Concurrently, KP activity toward a neuroprotective branch is reduced, which is characterized by kynurenic acid (KA), an NMDAR antagonist and anti-oxidant.^2,5^ The neurotoxic imbalance of kynurenines, expressed as an elevated QA/KA ratio, was demonstrated in the cerebrospinal fluid and serum of persons with MS compared to healthy individuals.^6^ A higher serum QA/KA ratio in persons with MS is related to higher disease severity,^7^ lower cognitive performance,^8^ and higher plasma neurofilament light chain (NfL) concentration,^8^ which is an established marker of neuroaxonal damage.^9^

These findings suggest that systemic kynurenines may reflect aspects of CNS pathophysiology in MS,^3^ underpinned by the fact that cerebrospinal fluid and systemic concentrations of kynurenines, including QA and KA, show moderate concordance.^7^ A narrow focus on QA and KA, or their (im)balance, however, does not capture the inherent complexity of the KP. An improved understanding of KP imbalance in MS requires holistic approaches beyond single metabolite investigations. This is especially relevant, as so far scarcely investigated kynurenines may also modulate the neurotoxic CNS microenvironment in MS, given their broad involvement in inflammatory processes, redox-regulation, and glutamatergic neurotransmission.^5^

Here we applied a state-of-the-art *targeted metabolomics* approach to determine serum concentrations of kynurenines in 353 persons with MS and 111 healthy individuals. Our objectives were to (1) validate MS-specific differences in the serum concentrations of kynurenines compared to healthy controls (HC), (2) explore KP metabolite patterns, and (3) assess their clinical relevance concerning disease severity, MS symptoms, magnetic resonance imaging (MRI) metrics, and relevant demographic, lifestyle-related, and disease-specific factors.

## Methods

### Study design and research ethics

This study is a pooled analysis of baseline data from eight randomized controlled trials conducted in Denmark, Germany, and Switzerland. Ethical approval for each study was obtained from the regional or institutional review board. All studies were conducted in accordance with the Declaration of Helsinki. Written informed consent was obtained from all participants.

### Study participants

The source studies included five MS cohorts (pooled MS cohort) and four HC cohorts (pooled HC cohort). Eligibility criteria for all studies were adult age (≥ 18 years) and the absence of severe acute or chronic disease states besides MS. Individual MS cohorts comprised participants of the EMSES (Denmark, NCT03322761), EXBRAIN (Denmark, NCT02661555), PoTOMS (Denmark, NCT04762342), HIITMS (Switzerland, NCT04356248), and FunTraiMS (Germany, DRKS00017091) studies. All persons with MS were diagnosed according to the McDonald criteria,^10,11^ had mild to moderate disease severity (Expanded Disability Status Scale (EDSS) score ≤ 6.5), and were stable on disease-modifying treatment, if any. Participants did not receive corticosteroid treatment at the time of blood sampling. Persons with relapsing-remitting MS (RRMS) were in the remission phase. Individual HC cohorts comprised matched healthy participants of the PoTOMS study and participants of the TRAIN (Germany, DRKS00031445), AhR (Germany, DRKS00028792), and METDEX (Germany, DRKS00029105) studies. The HIITMS study was conducted in an inpatient rehabilitation setting. All other studies were conducted in outpatient settings.

### Kynurenine pathway metabolite profiling

Blood samples were obtained from the antecubital vein after at least 5 min of rest. After clotting, samples were centrifuged at 1200 – 4300 g for 10 min. The supernatant was aliquoted and stored at -80°C at the study sites until analysis. Serum concentrations of kynurenines were analyzed in the same lab (Bevital AS, Bergen, Norway (https://bevital.no) between March 24, 2022 and August 9, 2024. Targeted metabolomics was performed using high-throughput liquid chromatography coupled with tandem mass spectrometry (LC-MS/MS).^12^ Serum concentrations of the following metabolites were determined: anthranilic acid (AA), KA, kynurenine (Kyn), neopterin (Neopt), picolinic acid (Pic), QA, quinaldic acid (Qld), Trp, xanthurenic acid (XA), 3-hydroxyanthranilic acid (3-HAA), and 3-hydroxykynurenine (3-HK). The assay’s lower limit of detection ranged from 0.01 to 8.00 nmol*L^-1^. Within-day coefficients of variation ranged from 3.78% to 9.32%.

### Assessment of clinical outcomes

The EDSS score was rated by neurologists. MRI metrics and data on walking capacity, fatigue, and cognitive performance were similarly collected in the EMSES, EXBRAIN, and PoTOMS studies, enabling data pooling. MRI scans were performed on the same 3-Tesla MRI scanner (MAGNETOM Skyra, Siemens Medical Systems, Erlangen, Germany). Structural T1-weighted (T1w) MP2RAGE images and T2-weighted fluid-attenuated inversion recovery (T2FLAIR) images were acquired. For this analysis, gray matter parenchymal fraction (GMPF), white matter parenchymal fraction (WMPF), total lesion volume, and volumes of the following regions of interest (ROI) were considered: hippocampus, thalamus, caudate, putamen, and globus pallidus. Walking capacity was assessed using the 6-Minute Walk Test (6MWT).^13^ Fatigue impact was queried using the 21-item Modified Fatigue Impact Scale (MFIS).^14,15^ Cognitive performance was tested using the Selective Reminding Test (SRT, verbal learning and memory) and the Symbol Digit Modalities Test (SDMT, information processing speed).^16,17^ Higher scores indicate greater fatigue and better cognitive performance, respectively. Body composition metrics included body mass index (BMI) and total body fat percentage. Body fat percentage was determined by bioimpedance analysis (BIA) in the EMSES and EXBRAIN study and by dual-energy x-ray absorptiometry (DXA) in the PoTOMS study. Cardiorespiratory fitness was assessed as peak oxygen consumption during cardiopulmonary exercise testing divided by body weight (relative V̇O_2 peak_) in all studies except the PoTOMS study.

### Statistical analysis

The workflow of statistical analyses is visualized in **Figure 1B**. KP metabolite data (i.e., serum concentrations of Neopt, Trp, and kynurenines) were screened for missing values, which were declared as “missing at random”. Data distribution was assessed visually using histograms. Due to relevant skewness, serum concentrations of all metabolites were uniformly log-transformed to approximate normal distribution. The KTR and QA/KA ratio were calculated subsequently.

In the first step, we assessed differences in serum concentrations of KP metabolites between the pooled MS and HC cohorts. We computed independent t-tests and adjusted the resulting p-values for false discovery rate (FDR).^18^ Subsequently, one-way ANCOVAs were computed to evaluate the influence of covariates, including sex, age, BMI, and sample storage time, on between-group differences in KP metabolite concentrations and ratios.

In the second step, we assessed correlations among KP metabolites and ratios separately for the MS and HC cohorts using Pearson’s correlation analysis. The results were refined using partial correlation analysis to account for potential confounding effects of sex, age, BMI, and relative V̇O_2 peak_. Correlations were FDR-adjusted and interpreted as weak (|*r*| = .10-.29), moderate (|*r*| = .30-.49), or strong (|*r*| ≥ .50).^19^ Data suitability for exploratory factor analysis (EFA) was checked comparing correlations and partial correlations and using the Kaiser-Meyer-Olkin (KMO) criterion. EFA was conducted using maximum likelihood estimation with promax rotation.^20^ The number of factors to retain was determined using the scree test and the Kaiser-Guttmann criterion. The EFA-derived factors were designated according to current evidence on the neuroactive properties ascribed to the KP metabolites and ratios with relevant loadings (|λ| ≥ .3) to characterize KP metabolite patterns. Trp, the precursor metabolite, and metabolites with relevant loadings on more than one extracted factor (i.e., 3-HAA) were neglected in pattern designation. Consistency of KP metabolite patterns across individual MS cohorts was checked using EFAs with a predefined two-factor solution. Using the same approach, we additionally explored the presence of similar KP metabolite patterns in the HC cohorts.

In the final third step, we investigated associations of the KP metabolite patterns with clinical outcomes. Correlation analyses were repeated to assess associations between the KP metabolite patterns and demographic (i.e., age), anthropometric (i.e., BMI, body fat percentage), and disease-related characteristics (i.e., EDSS score, serum NfL concentration, relative V̇O_2 peak_). Associations of the KP metabolite patterns with EDSS score were computed using a logistic proportional odds regression model. Associations of the KP metabolite patterns with MRI metrics and MS symptoms were assessed using multiple linear regression models (MLRs). MLRs included the KP metabolite patterns, sex, age, BMI, EDSS score, disease-modifying treatment (yes/no), total lesion volume, and study (EMSES, EXBRAIN, PoTOMS) as predictors, and 6MWT distance, MFIS total score, SRT and SDMT scores, and MRI metrics as predicted variables. In all analyses, the significance level was set at p= .05. Statistical analyses were conducted using R (version 4.4.2).^21^ Figures were created with R and biorender.com.

## Results

### Cohort characteristics

Characteristics of the pooled MS and HC cohorts, along with individual subcohorts, are presented in **Figure 1C**. Disease-specific characteristics of the MS cohorts, both pooled and individual, are shown in **Table 1**. In brief, the pooled MS cohort included 353 participants (67.1% female) with a mean (SD) age of 46.1 (12.4) years. Most participants had the RRMS phenotype (74.5%) and were receiving disease-modifying treatment (74.2%). The mean (SD) EDSS score was 3.1 (1.8). The pooled HC cohort included 111 participants (53.2% female), with a mean (SD) age of 45.7 (16.6) years.

**Table 1.**
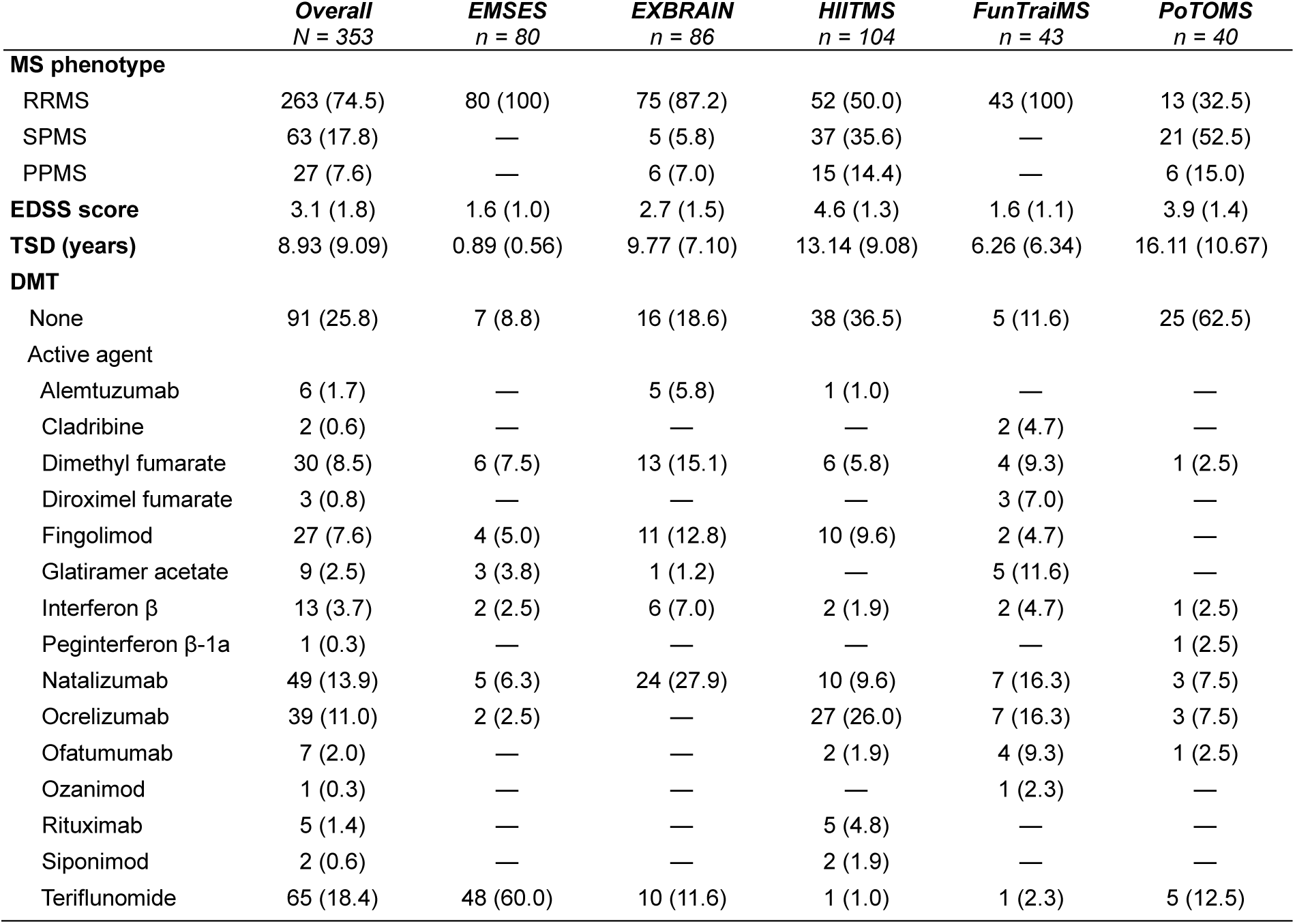
Disease-related characteristics of the pooled MS cohort along with subcohorts. Categorical data are reported as the total number and percentage (%). Continuous data are expressed as mean (SD). Missing data include information on disease-modifying treatment (n = 3) and Expanded Disability Status Scale scores (n = 9) for individual participants in the EMSES study. Time since diagnosis was missing for one participant in the FunTraiMS study. DMT = disease-modifying treatment; EDSS = Expanded Disability Status Scale; MS = multiple sclerosis; PPMS = primary progressive MS; RRMS = relapsing-remitting multiple sclerosis; SPMS= secondary progressive multiple sclerosis; TSD = time since diagnosis.

|  | *Overall*<br>*N = 353* | *EMSES*<br>*n = 80* | *EXBRAIN*<br>*n = 86* | *HIITMS*<br>*n = 104* | *FunTraiMS*<br>*n = 43* | *PoTOMS*<br>*n = 40* |
| --- | --- | --- | --- | --- | --- | --- |
| **MS phenotype** |  |  |  |  |  |  |
| RRMS | 263 (74.5) | 80 (100) | 75 (87.2) | 52 (50.0) | 43 (100) | 13 (32.5) |
| SPMS | 63 (17.8) | — | 5 (5.8) | 37 (35.6) | — | 21 (52.5) |
| PPMS | 27 (7.6) | — | 6 (7.0) | 15 (14.4) | — | 6 (15.0) |
| **EDSS score** | 3.1 (1.8) | 1.6 (1.0) | 2.7 (1.5) | 4.6 (1.3) | 1.6 (1.1) | 3.9 (1.4) |
| **TSD (years)** | 8.93 (9.09) | 0.89 (0.56) | 9.77 (7.10) | 13.14 (9.08) | 6.26 (6.34) | 16.11 (10.67) |
| **DMT** |  |  |  |  |  |  |
| None | 91 (25.8) | 7 (8.8) | 16 (18.6) | 38 (36.5) | 5 (11.6) | 25 (62.5) |
| Active agent |  |  |  |  |  |  |
| Alemtuzumab | 6 (1.7) | — | 5 (5.8) | 1 (1.0) | — | — |
| Cladribine | 2 (0.6) | — | — | — | 2 (4.7) | — |
| Dimethyl fumarate | 30 (8.5) | 6 (7.5) | 13 (15.1) | 6 (5.8) | 4 (9.3) | 1 (2.5) |
| Diroximel fumarate | 3 (0.8) | — | — | — | 3 (7.0) | — |
| Fingolimod | 27 (7.6) | 4 (5.0) | 11 (12.8) | 10 (9.6) | 2 (4.7) | — |
| Glatiramer acetate | 9 (2.5) | 3 (3.8) | 1 (1.2) | — | 5 (11.6) | — |
| Interferon $\beta$ | 13 (3.7) | 2 (2.5) | 6 (7.0) | 2 (1.9) | 2 (4.7) | 1 (2.5) |
| Peginterferon β-1a | 1 (0.3) | — | — | — | — | 1 (2.5) |
| Natalizumab | 49 (13.9) | 5 (6.3) | 24 (27.9) | 10 (9.6) | 7 (16.3) | 3 (7.5) |
| Ocrelizumab | 39 (11.0) | 2 (2.5) | — | 27 (26.0) | 7 (16.3) | 3 (7.5) |
| Ofatumumab | 7 (2.0) | — | — | 2 (1.9) | 4 (9.3) | 1 (2.5) |
| Ozanimod | 1 (0.3) | — | — | — | 1 (2.3) | — |
| Rituximab | 5 (1.4) | — | — | 5 (4.8) | — | — |
| Siponimod | 2 (0.6) | — | — | 2 (1.9) | — | — |
| Teriflunomide | 65 (18.4) | 48 (60.0) | 10 (11.6) | 1 (1.0) | 1 (2.3) | 5 (12.5) |

### Serum concentrations of kynurenine pathway metabolites and ratios differ between persons with MS and healthy individuals

The KTR (**Figure 2D**), the serum concentration of Trp (**Figure 2B**), and serum concentrations of most kynurenines were lower in the pooled MS cohort compared to the HC cohort. These kynurenines included Kyn (**Figure 2C**), KA (**Figure 2E**), AA (**Figure 2F**), XA (**Figure 2G**), Pic (**Figure 2I**), and QA (**Figure 2J**). Serum concentrations of the inflammation marker Neopt (**Figure 2A**) and 3-HAA (**Figure 2H**), and the QA/KA ratio (**Figure 2K**) were higher in the pooled MS cohort. Serum concentrations of Qld and 3-HK did not differ between cohorts (data not shown).

**Figure 2.**
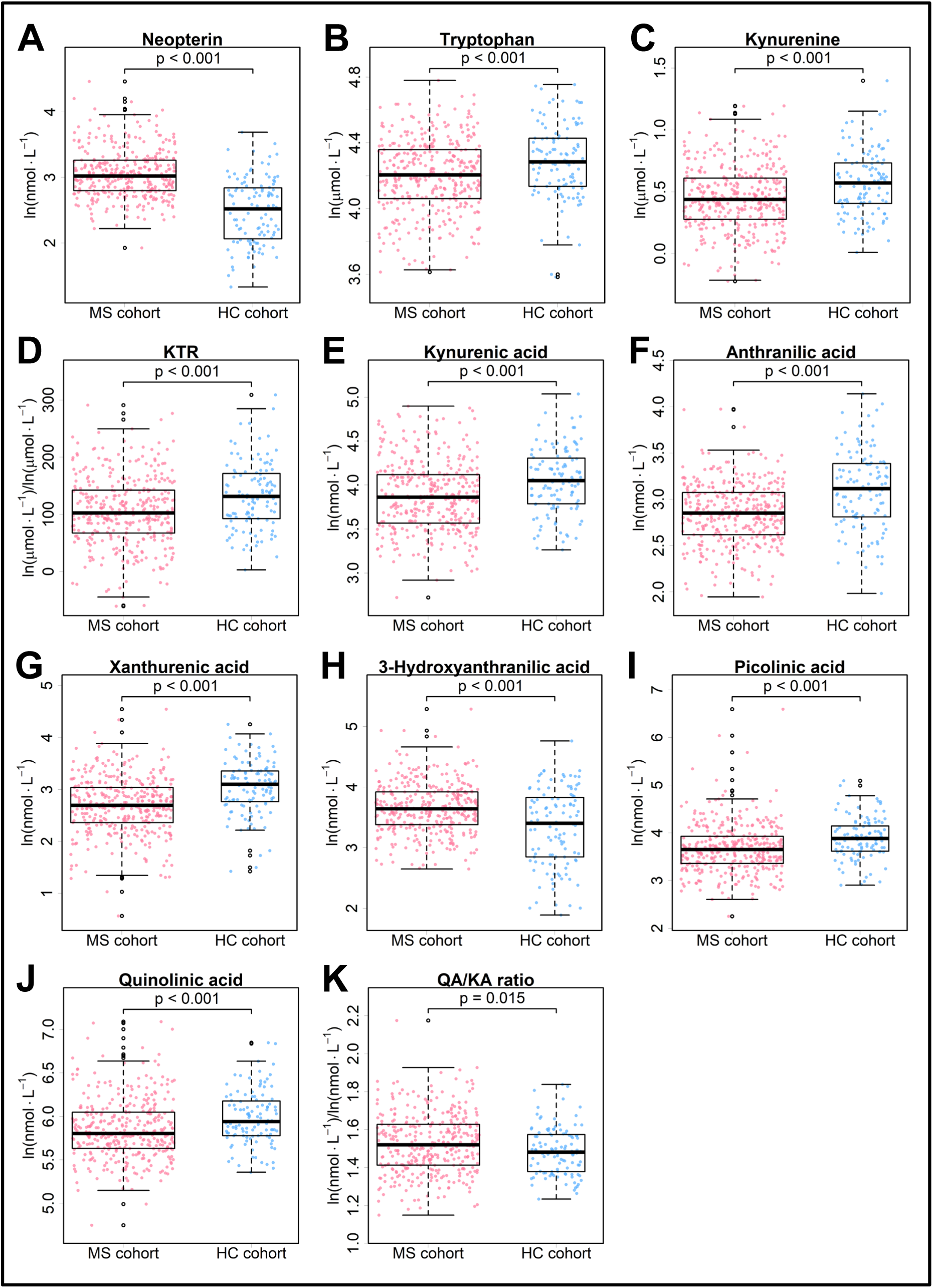
Differences in serum concentrations of kynurenine pathway metabolites and ratios between the pooled MS and HC cohorts. Box plots illustrating significant differences in log-transformed serum concentrations of neopterin, tryptophan, kynurenines, and metabolite ratios between the pooled multiple sclerosis (red) and healthy cohorts (blue). No between-group differences were found for quinaldic acid and 3-hydroxykynurenine (not shown). P-values refer to the results of independent t-tests and were adjusted for false discovery rate. HC = healthy cohort; KTR = kynurenine-to-tryptophan ratio; MS = multiple sclerosis; QA/KA ratio = quinolinic acid-to-kynurenic acid ratio.

### Correlation matrices differ between persons with MS and healthy individuals

Visual inspection of the correlation matrices revealed marked differences in the strength and direction of correlations among KP metabolites and ratios between the pooled MS and HC cohorts (**Figure 1D**). For example, a higher Neopt concentration was correlated with a higher QA/KA ratio only in the MS cohort. Additionally, Neopt showed stronger positive correlations with KTR, 3-HK concentration, and QA concentration in the MS cohort compared to the HC cohort.

We also identified some MS-specific differences in correlations between KP metabolites and ratios and lifestyle-related factors. For example, a higher BMI was correlated with a higher KTR and higher concentrations of most KP metabolites, including Neopt, Kyn, KA, 3-HK, XA, 3-HAA, and QA, in the pooled MS cohort but not in the HC cohort.

Overall, these correlations suggest that, in persons with MS, kynurenines have stronger associations with inflammation, indicated by the inflammation marker Neopt, and lifestyle-related factors influencing the inflammatory microenvironment.

### Persons with MS reveal two distinct kynurenine pathway metabolite patterns: “NeuroTox” and “NeuroPro”

Correlations among KP metabolites, which were not simply explained by bivariate correlations, and a KMO criterion of 0.715 (middling) in the pooled MS cohort indicated data suitability for EFA. Application of the Kaiser-Guttmann criterion and the scree test resulted in a two-factor solution (**Figure 3A**). The cumulative proportional variance explained by the two factors was 54.2%.

**Figure 3.**
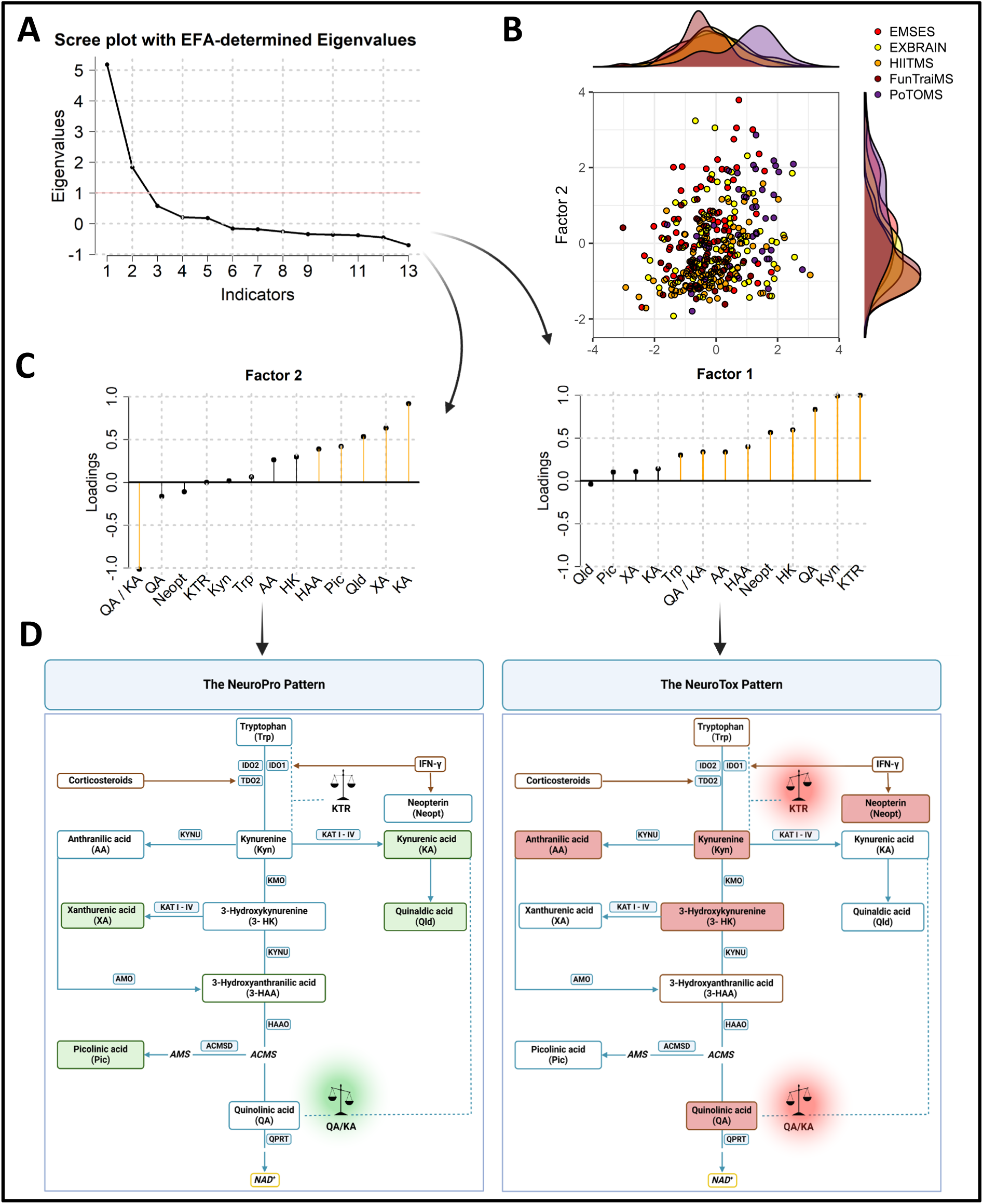
The pooled MS cohort reveals two distinct kynurenine pathway metabolite patterns. **A** Scree plot illustrating the number of factors to be retained for exploratory factor analysis (EFA). **B** Combined score and density plot illustrating the EFA-derived factors of participants, colored by source cohort (score plot), and the distribution of both factors at the cohort level (density plot). **C** Lollipop plots showing the loadings of individual kynurenine pathway metabolites and ratios on the two EFA-derived factors. Relevant loadings (|λ| ≥ .3) are highlighted in yellow. **D** Representation of the EFA-derived factors as distinct serum kynurenine pathway metabolite patterns according to Figure 1A. Factor 1 represents an inflammation-driven neurotoxic kynurenine pathway metabolite pattern and is referred to as “NeuroTox”. Factor 2 represents a neuroprotective kynurenine pathway metabolite pattern and is referred to as “NeuroPro”. As a precursor metabolite, tryptophan was not considered in factor designation of “NeuroTox”. 3-hydroxyanthranilic acid showed relevant loadings on both factors and was therefore neglected in factor designation. AA = anthranilic acid; KA = kynurenic acid; KTR = kynurenine-to-tryptophan ratio; Kyn = kynurenine; Neopt = neopterin; Pic = picolinic acid; Trp = tryptophan; QA = quinolinic acid; QA/KA ratio = quinolinic acid-to-kynurenic acid ratio; Qld = quinaldic acid; XA = xanthurenic acid; 3-HAA = 3-hydroxyanthranilic acid; 3-HK = 3-hydroxykynurenine.

Factor 1 and Factor 2 were differentially distributed among participants as a function of source cohort. Factor 1 was particularly pronounced in the PoTOMS cohort, while it was less pronounced and more homogeneously distributed in the EMSES cohort. Factor 2 was less pronounced and more homogeneously distributed in the EMSES and PoTOMS cohorts (**Figure 3B)**.

Factor 1 was characterized by positive loadings of the inflammation marker Neopt and the KTR, indicating inflammatory stimulation of KP activity. Factor 1 was also characterized by positive loadings of the predominantly neurotoxic kynurenines 3-HK and QA, alongside the neurotoxicity index, the QA/KA ratio (**Figure 3C, right**). Thus, Factor 1 was considered to reflect an inflammation-driven neurotoxic KP metabolite pattern, and hereafter referred to as “NeuroTox” (**Figure 3D, right**).

Factor 2 was characterized by positive loadings of the neuroprotective KP metabolite KA, its degradation product quinaldic acid (Qld), xanthurenic acid (XA), and picolinic acid (Pic), and an inverse loading of the QA/KA ratio (**Figure 3C, left**). Factor 2 was thus designated as a neuroprotective KP metabolite pattern, and referred to as “NeuroPro” (**Figure 3D, left**).

Translating the emergent KP metabolite patterns to the individual MS cohorts revealed consistent loadings of KP metabolites and ratios on “NeuroTox" and “NeuroPro". In all MS cohorts, “NeuroTox" was characterized by positive loadings of th KTR, Kyn, 3-HK, and QA, whereas “NeuroPro" was characterized by positive loadings of KA and XA, and an inverse loading of the QA/KA ratio (**Figure 4**,

**Figure 4.**
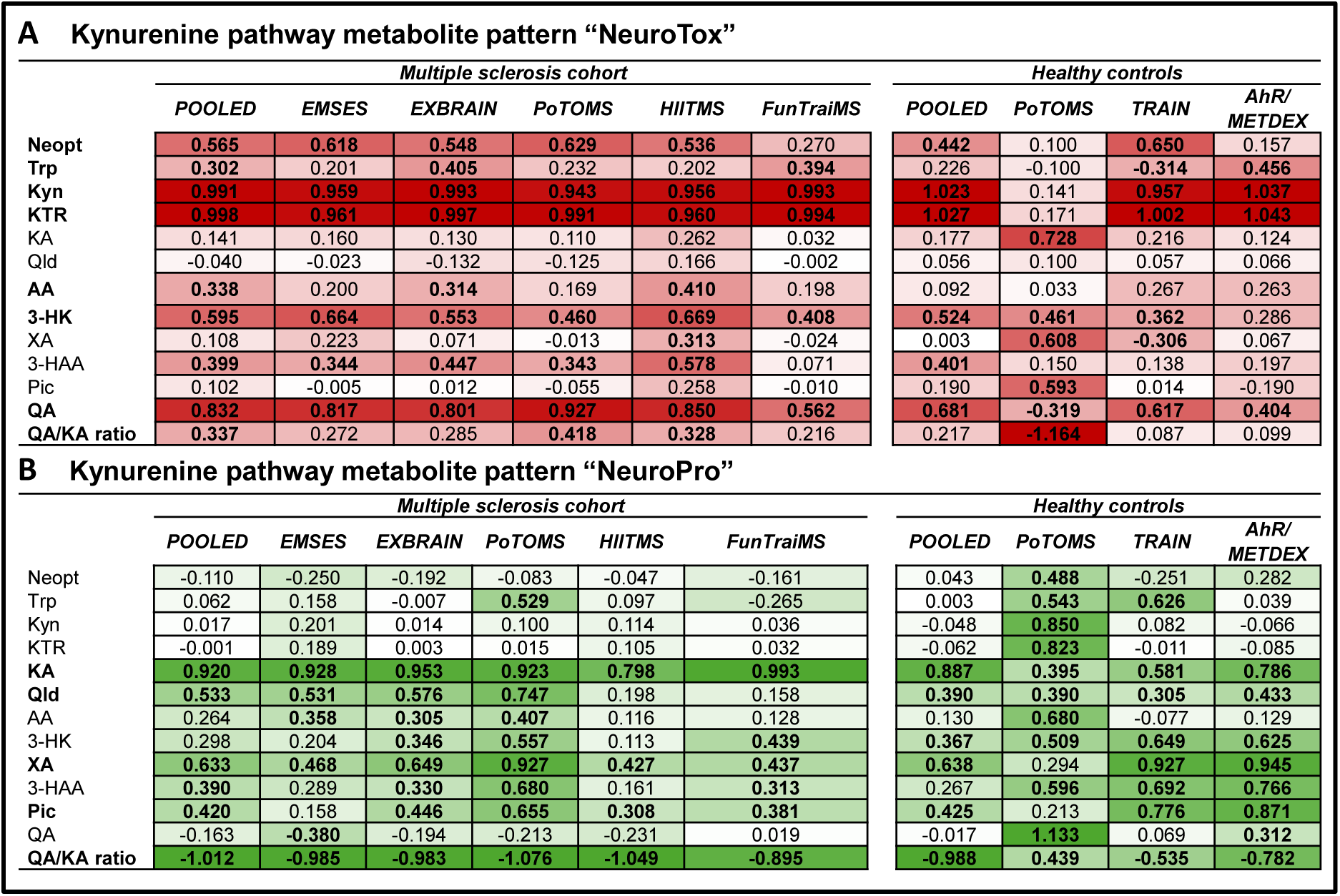
Loadings of kynurenine pathway metabolites and ratios on the two EFA-derived factors in individual MS and HC cohorts. Heatmaps illustrating the similarities and differences in kynurenine pathway metabolite patterns across individual multiple sclerosis cohorts and between the pooled multiple sclerosis and healthy cohorts. **A** Loadings on Factor 1 referred to as the inflammation-driven kynurenine pathway metabolite pattern “NeuroTox”. **B** Loadings on Factor 2 referred to as the neuroprotective kynurenine pathway metabolite pattern “NeuroPro”. Darker color indicates stronger positive or inverse loadings of the kynurenine pathway metabolite or ratio on the respective factor. In the left column, kynurenine pathway metabolites and ratios with relevant loadings (|λ| ≥ .3) in the main analysis (Figure 3C) are printed in bold. AA = anthranilic acid; KA = kynurenic acid; KTR = kynurenine-to-tryptophan ratio; Kyn = kynurenine; Neopt = neopterin; Pic = picolinic acid; Trp = tryptophan; QA = quinolinic acid; QA/KA ratio = quinolinic acid-to-kynurenic acid ratio; Qld = quinaldic acid; XA = xanthurenic acid; 3-HAA = 3-hydroxyanthranilic acid; 3-HK = 3-hydroxykynurenine.

**left**). Taking into account limited translatability, we exploratively assessed the presence of similar KP metabolite patterns in the pooled HC cohort. The pooled HC cohort showed some similarities in the loadings of KP metabolites and ratios on both predefined factors, such as positive loadings of Neopt, KTR, Kyn, 3-HK, and QA on “NeuroTox", and positive loadings of KA, Qld, XA, and Pic together with an inverse loading of the QA/KA ratio on “NeuroPro". However, in comparison to the pooled MS cohort, “NeuroTox" revealed reduced loading of QA and no relevant (positive) loadings of AA and the QA/KA ratio (**Figure 4, right**).

### Kynurenine pathway metabolite patterns are associated with clinical outcomes

Greater “NeuroTox” (**Figure 5A**) and lower “NeuroPro” (**Figure 5B**) were correlated with higher disease severity (EDSS score). Greater “NeuroTox” was also correlated with older age (**Figure 5C**), higher BMI (**Figure 5D**), and higher body fat percentage, as determined by BIA (**Figure 5E**) and DXA (**Figure 5F**). “NeuroPro” was associated with lower body fat percentage (**Figure 5G**) and higher cardiorespiratory fitness (relative V̇O_2 peak_) (**Figure 5H**). No correlations were observed between KP metabolite patterns and serum NfL concentration (EXBRAIN cohort) (**Suppl. Figure 1**).

**Figure 5.**
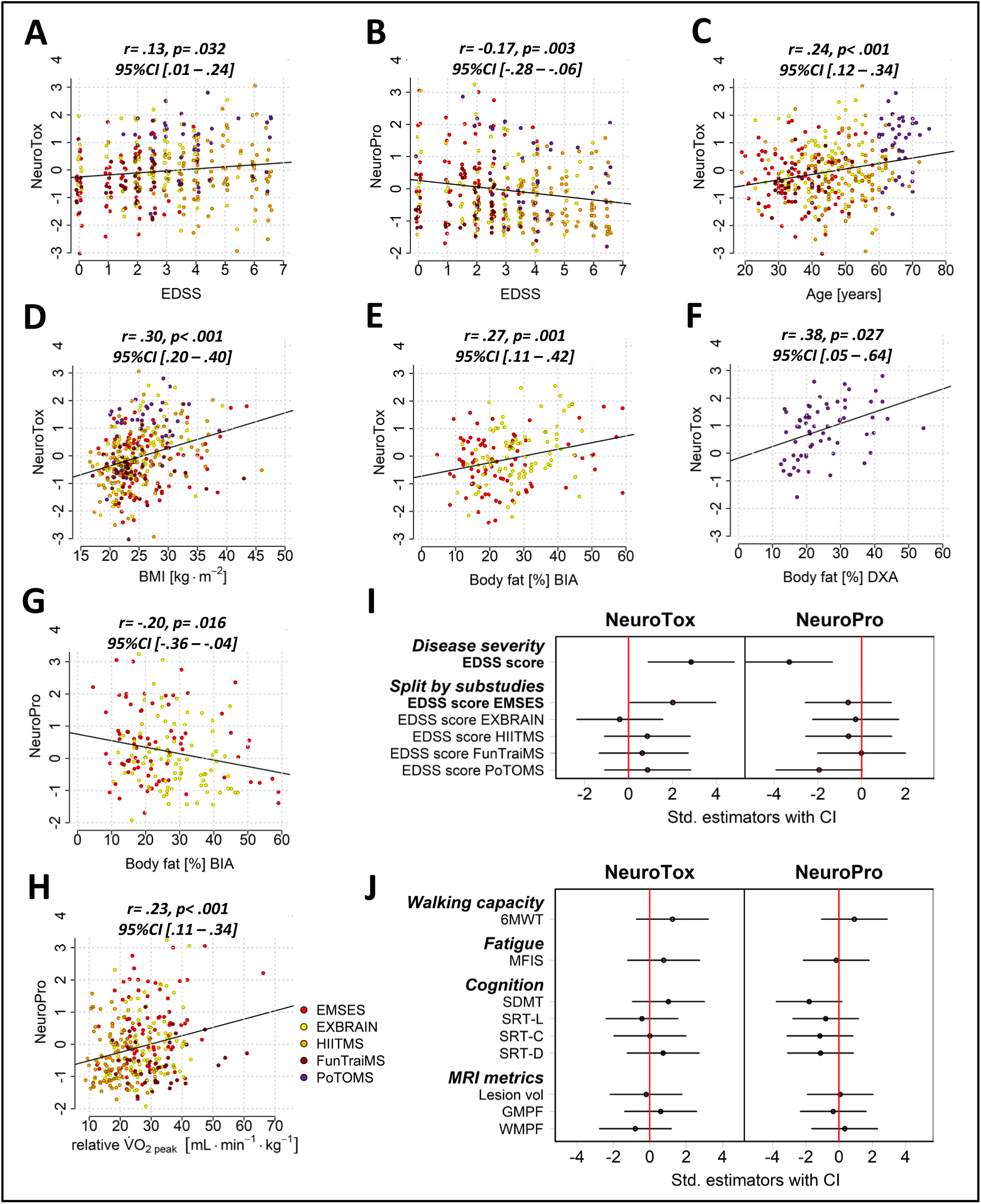
Associations between the kynurenine pathway metabolite patterns “NeuroTox” and “NeuroPro” with participant characteristics, MS symptoms, and MRI metrics in the pooled MS cohort and subcohorts. **A-H** Scatter plots showing correlations between “NeuroTox” and “NeuroPro” with disease severity (EDSS score), age, and body mass index including all MS cohorts, cardiorespiratory fitness (relative V̇O_2 peak_) including all MS cohorts except PoTOMS, and total body fat percentage, determined by either bioimpedance analysis (EMSES and EXBRAIN) or dual-energy x-ray absorptiometry (PoTOMS). P-values were adjusted for false discovery rate. **I** Estimator plots showing the standardized estimators of the proportional odds regression models with “NeuroTox” and “NeuroPro” as predictor variables and disease severity (EDSS score) as predicted variable. Regression analyses were performed based on data from the pooled MS cohort (EMSES, EXBRAIN, HIITMS, FunTraiMS, and PoTOMS) and individual cohorts. Greater “NeuroTox” was associated with higher disease severity in the EMSES cohort (bold). **J** Estimator plots showing the standardized estimators of the multiple linear regression models with “NeuroTox” and “NeuroPro” as predictor variables and walking capacity (6MWT), fatigue (MFIS), cognitive performance (SDMT, SRT), or MRI metrics (total lesion volume, GMPF, WMPF) as predicted variables. Regression analyses were performed based on data from the pooled EMSES, EXBRAIN, and PoTOMS cohorts. BIA = bioimpedance analysis; BMI = body mass index; DXA = dual-energy x-ray absorptiometry; EDSS = Expanded Disability Status Scale; GMPF = gray matter parenchymal fraction; MFIS = Modified Fatigue Impact Scale (total score); SDMT = Symbol Digit Modalities Test; SRT= Selective Reminding Test; SRT-L = Selective Reminding Test-Long-term Retrieval Test; SRT-C = Selective Reminding Test-Consistent Long-term Retrieval Test; SRT-D = Selective Reminding Test-Delayed Recall Test; WMPF = white matter parenchymal fraction; relative V̇O_2 peak_ = peak oxygen consumption during cardiopulmonary exercise testing divided by kilograms body weight; 6MWT = Six-Minute Walk Test.

The standardized estimates of the proportional odds regression models indicate a trend toward associations between greater “NeuroTox” and lower “NeuroPro” and higher EDSS score. When considering individual cohorts, we showed that greater “NeuroTox” was significantly associated with higher EDSS score in the EMSES cohort (**Figure 5I**). “NeuroTox” and “NeuroPro” were not associated with walking capacity (6MWT), fatigue (MFIS), cognitive performance (SDMT, SRT), or MRI metrics (**Figure 5J, Suppl. Figure 2**).

## Discussion

In this pooled analysis, we determined serum concentrations of kynurenines in a large cohort of 353 persons with MS and 111 healthy individuals. Firstly, we investigated MS-specific differences in serum concentrations of KP metabolites and ratios compared to healthy individuals. Secondly, we explored KP metabolite patterns in persons with MS, which were then evaluated in terms of consistency across individual MS cohorts and compared to KP metabolite patterns in healthy individuals. Thirdly, we investigated associations between the emergent KP metabolite patterns in the pooled MS cohort, alongside individual kynurenines and metabolite ratios, with participant characteristics and clinical outcomes to study the role of the KP as one of the neuroimmunological pathways involved in MS pathophysiology.

We show that persons with MS had a lower KTR and lower concentrations of most kynurenines compared to healthy individuals. These kynurenines include both metabolites ascribed with neurotoxic and those ascribed with neuroprotective properties. The serum concentration of Neopt and the QA/KA ratio were higher in persons with MS (**Figure 2A, 2K**). Neopt is thought to indicate Th1-mediated inflammatory stimulation of the KP.^22^ Therefore, higher systemic concentrations of kynurenines in healthy individuals may result from inflammation-independent mechanisms. It may be hypothesized that persons with MS, in general presenting with a lower physical activity level^23^ and reduced cardiorespiratory fitness^24^ compared to healthy individuals, have a lower bioenergetic turnover to generate NAD^+^, considering that both the hepatic and skeletal muscle KP substantially contribute to systemic concentrations of kynurenines.^25,26^ In line with that, we recently discussed exercise-induced KP modulation in persons with MS as a potential endogenous mechanism to cover increased NAD^+^ demands.^27^ Additionally, it has also been shown that persons with MS exhibit reduced expression of the KP enzyme indoleamine 2,3-dioxygenase 1 (IDO1) in peripheral blood mononuclear cells.^28^ As IDO1 induces the first step of the KP,^26^ reduced IDO1 expression in immune cells may contribute to lower concentrations of kynurenines in the systemic circulation.

Earlier studies yielded discrepant results concerning differences in systemic concentrations of kynurenines between persons with MS and healthy individuals, challenging comparisons with our findings. Nevertheless, we replicate the most consistent finding of human MS trials, that is a higher QA/KA ratio in persons with MS.^29,30^

Using a data-driven approach, we identified two distinct KP metabolite patterns (**Figure 3**) that were consistent across individual MS studies (**Figure 4**). These two patterns separated neurotoxic and neuroprotective kynurenines, together with the respective precursors and degradation products, according to the traditional view of a neurotoxic and neuroprotective KP branch.^2^ Beyond that, our results highlight that kynurenines, with as yet unclear roles in MS, also clearly allocated to either “NeuroTox” or “NeuroPro”.

The emergent KP metabolite patterns indicate that persons with pronounced “NeuroTox” present with elevated systemic inflammation, increased KP activity, and neurotoxic KP imbalance toward the formation of the neurotoxin QA. “NeuroTox” also included 3-HK, another neurotoxin, pro-oxidant, and QA-agonist.^3^ In comparison, persons with pronounced “NeuroPro” may have an increased neuroprotective capacity, considering that “NeuroPro” was characterized by the neuroprotective and anti-oxidative metabolite KA^2,5^ and its precursor Qld, in line with a shift of the QA/KA ratio toward KA. “NeuroPro” also included XA and Pic. Albeit ascribed Janus-faced roles, both kynurenines have been shown to attenuate aspects of QA-related neurotoxicity and may thus agonize the neuroprotective effects of KA.^3^

Given the concordance of cerebrospinal fluid and systemic concentrations of kynurenines,^7^ both an elevated QA/KA ratio in persons with MS and a shift in KP metabolite patterns may mirror KP imbalance at the CNS level and its potential contribution to the neurotoxic CNS microenvironment inherent to MS. Accordingly, persons with pronounced “NeuroTox” and/or reduced “NeuroPro” would be particularly vulnerable to neuroaxonal damage. We do not show associations between KP metabolite patterns and MRI metrics or MS symptoms, which might be explained by the investigation of a rather well-functioning study cohort (mean (SD) EDSS score of 3.1 (1.8)) that likely does not show extensive structural brain damage, walking impairment, fatigue, or cognitive impairment. This aspect has been previously discussed in relation to a low prevalence of cognitive impairment in the EMSES and EXBRAIN cohorts.^31,32^ Nevertheless, we demonstrate that a greater “NeuroTox” and its components, Neopt, KTR, QA/KA ratio, and QA, correlated with higher disease severity (**Figure 5A, Suppl. Figure 1**).

Concordantly, a greater “NeuroPro” and its components, KA and XA, correlated with lower disease severity (**Figure 5B, Suppl. Figure 1**). While the same trend was reproduced in the logistic regression models, the associations remained nonsignificant in the adjusted analyses, potentially due to the significant association between age and EDSS score. In support of this notion, both greater “NeuroTox” and lower “NeuroPro” were significantly associated with higher EDSS score when excluding the covariates age and study cohort from the models.

Aging and MS are thought to induce a cumulative impact on neuroaxonal damage in older persons with MS via the interaction of age- and disease-specific mechanisms that are detrimental to CNS health.^33^ These mechanisms likely involve the KP, given that immunological changes during the aging process, referred to as inflammaging, are themselves associated with KP imbalance.^34^ In that sense, inflammaging contributes to systemic KP imbalance due to the perpetuation of chronic systemic low-grade inflammation, and may fuel CNS KP imbalance due to an increased activation of microglia and CNS recruitment of pro-inflammatory macrophages,^33^ both of which are the main source of QA in the CNS.^2^ In line with that, we show that older age was associated with greater "NeuroTox" (**Figure 5C**) and that participants of the oldest cohort PoTOMS (mean (SD) age: 66.7 (4.0) years) revealed particularly pronounced “NeuroTox”, especially when compared to the youngest cohort EMSES (mean (SD) age: 37.3 (9.9) years) (**Figure 1C**, **Figure 3B** (density plot)).

Our findings additionally suggest that both “NeuroTox” and “NeuroPro” are modulated by lifestyle-related factors, such as body composition and regular physical exercise, both of which shape the pro-inflammatory microenvironment in MS and influence MS-related disability.

In this regard, it has been shown that hypertrophic and dysfunctional adipose tissue contributes to chronic systemic low-grade inflammation via the release of pro-inflammatory cytokines, such as interferon-γ, thereby contributing to increased inflammatory stimulation of the KP.^35^ Accordingly, we show that a higher BMI and body fat percentage were associated with greater “NeuroTox” across measurement methods (**Figure 5D-F**). These findings confirm our previous results showing that overweight and obese persons with MS had a higher KTR and higher serum concentrations of most kynurenines compared to normal weight and underweight persons with MS.^36^ Expanding our previous results, we now demonstrate that a higher body fat percentage is associated with a predominant formation of neurotoxic metabolites, such as QA. These findings are of particular relevance for the potential involvement of the KP in MS pathophysiology, considering that obesity in childhood and adolescence significantly increases MS risk,^37^ and that obese persons with MS exhibit higher disease severity and accelerated disability accumulation.^38^

In contrast, regular physical exercise, that is indicated by increased cardiorespiratory fitness, has a beneficial impact on the pro-inflammatory microenvironment in MS and KP imbalance. We previously showed that exercise reduced systemic markers of inflammation^39^ and modulated the protein expression in CD8^+^ T cells, a lymphocyte population targeted by natalizumab, a well-established disease-modifying treatment for MS.^40^ Moreover, we previously reported that an acute intense exercise bout shifted the QA/KA ratio toward an increased formation of KA.^8^ Results of this study show that higher cardiorespiratory fitness was associated with greater “NeuroPro” (**Figure 5H**) and its components, including KA, Qld, XA, Pic, and QA/KA ratio (inversely) (**Figure 1D**). KA, XA, and Pic may induce a concerted neuroprotective and/or anti-inflammatory action, given that, for example, both KA and XA are ligands to the aryl hydrocarbon receptor,^41^ which promotes the differentiation of anti-inflammatory regulatory T cells.^42^ In this regard, laquinimod, another aryl hydrocarbon receptor ligand with structural similarity to KA, has been shown to induce potent disease-modifying effects in two phase III trials in person with MS.^43^

Our study comes with strengths and limitations. To our knowledge, this study is the largest investigation of the KP in MS. Included participants are well characterized in terms of demographic, anthropometric, and disease-specific characteristics, and cover a wide age (19 – 76 years) and disability (EDSS ≤ 6.5) range. Targeted metabolomics analyses of MS and healthy cohorts have been performed in the same lab using state-of-the-art LC-MS/MS methodology to determine serum concentrations of the *complete* set of kynurenines. By applying a holistic data-driven approach that, beyond single metabolites investigations, enabled the identification of distinct KP metabolite patterns, we accounted for the complexity of the KP and the interrelation of kynurenines. Using the KP metabolite patterns “NeuroTox” and “NeuroPro”, we comprehensively investigated associations between the systemic KP and markers of neuroaxonal damage (e.g., MRI metrics), disease severity, MS symptoms, and a broad collection of MS-relevant participant-specific data. Thereby, the results of this analysis improve the current understanding of the KP and its potential clinical relevance in MS. However, as we pooled baseline data from exercise trials, mostly conducted in ambulatory settings, our study population may be affected by selection bias in terms of a high number of participants that are functionally independent and follow a health-promoting lifestyle, thus limiting the generalizability of the results. Additional control for diet, supplement intake, and MS-specific factors, such as time since the last relapse, would have been optimal. Inherent to the cross-sectional analysis of systemic kynurenines, our results do not allow statements on the cellular or tissue-specific origin of kynurenines, intercompartmental exchange across the blood-brain barrier, or causality of associations. In this regard, future studies are warranted that include participants across all disability levels, perform repeated blood sampling, and include additional outcomes, such as comprehensive immunophenotyping.

## Conclusion

This study shows that persons with MS reveal lower serum concentrations of most kynurenines, but a higher concentration of the inflammation marker Neopt and a higher QA/KA ratio compared to healthy individuals. Using a novel data-driven approach that accounts for the complexity of the KP, we identified two distinct KP metabolite patterns in persons with MS that resemble and expand the traditional view of a neurotoxic and neuroprotective KP branch: “NeuroTox” (Neopt, KTR, Kyn, 3-HK, AA, QA, QA/KA ratio) and “NeuroPro” (KA, Qld, XA, Pic, inverse QA/KA ratio). The KP metabolite patterns were not associated with MRI metrics or MS symptoms in our well-functioning study population. However, greater “NeuroTox” and lower “NeuroPro” were associated with higher disease severity. Our findings additionally indicate that age and lifestyle-related factors, such as body composition and regular physical exercise, interact with KP (im)balance, with potential implications for MS pathophysiology that require further study. Future studies may use comparable data-driven approaches to investigate whether KP imbalance follows similar disease-specific patterns in diseases other than MS.

## Data Availability

Data that support the findings of this study are available from the corresponding author upon reasonable request.

## Author contributions

Marie Kupjetz: Conceptualization, Formal analysis, Investigation, Funding Acquisition, Methodology, Visualization, Writing – original draft; Martin Langeskov-Christensen: Investigation, Funding Acquisition, Project administration, Resources, Writing – review & editing; Morten Riemenschneider: Investigation, Funding Acquisition, Project administration, Resources, Writing – review & editing; Stefan Inerle: Data curation, Software, Formal analysis, Methodology, Visualization, Writing – review & editing; Uwe Ligges: Data curation, Software, Formal Analysis, Methodology, Visualization, Writing – review & editing; Tobias Gaemelke: Investigation, Resources, Funding Acquisition, Writing – review & editing; Nadine Patt: Investigation, Resources, Writing – review & editing; Jens Bansi: Investigation, Funding Acquisition, Project administration, Resources, Writing – review & editing; Roman Gonzenbach: Writing – review & editing; Marcel Reuter: Investigation, Resources, Writing – review & editing; Friederike Rosenberger: Investigation, Resources, Writing – review & editing; Meyer, T.: Writing – review & editing; Adrian McCann: Resources, Writing – review & editing; Per Magne Ueland: Resources, Writing – review & editing; Simon Fristed Eskildsen: Data curation, Resources, Software, Writing – review & editing; Nygaard, Mikkel Karl Emil Nygaard: Data curation, Resources, Software, Writing – review & editing; Niklas Joisten: Conceptualization, Writing – review & editing; Lars Grøndahl Hvid: Funding Acquisition, Project administration, Writing – review & editing; Ulrik Dalgas: Funding Acquisition, Project administration, Writing – review & editing; Philipp Zimmer: Conceptualization, Funding Acquisition, Project administration, Supervision, Writing – review & editing.

## Figures

**Suppl. Figure 1.**
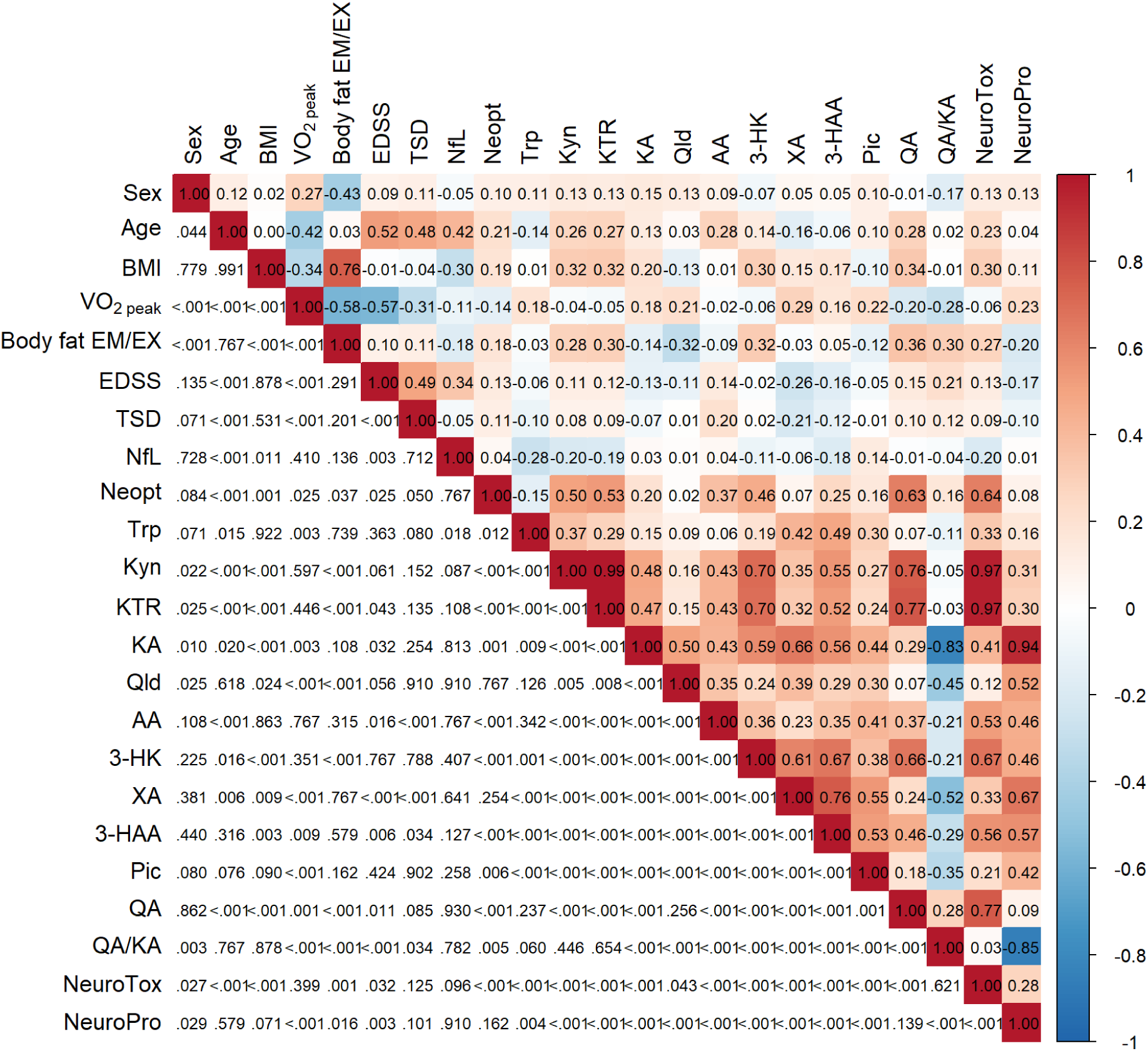
Heatmap showing correlations between kynurenine pathway metabolite patterns, participant characteristics, disease-related factors, and individual kynurenines and ratios. Correlations were calculated using two-tailed Pearson’s correlation analysis. The lower triangle of the heatmap shows false discovery rate-adjusted p-values. The upper triangle of the heatmaps show Pearson’s r correlation coefficients. Body fat was obtained using bioimpedance analysis in the EMSES and EXBRAIN study. AA = anthranilic acid; BMI = body mass index; EDSS = Expanded Disability Status Scale; KA = kynurenic acid; KTR = kynurenine-to-tryptophan ratio; Kyn = kynurenine; Neopt = neopterin; NfL= neurofilament light chain (determined in serum); Pic = picolinic acid; Trp = tryptophan; TSD = time since diagnosis (highest available precision, years or months); QA = quinolinic acid; QA/KA = quinolinic acid-to-kynurenic acid ratio; Qld = quinaldic acid; V̇O_2 peak_ = peak oxygen consumption during cardiopulmonary exercise testing divided by kilograms body weight; XA = xanthurenic acid; 3-HAA = 3-hydroxyanthranilic acid; 3-HK = 3-hydroxykynurenine.

**Suppl. Figure 2.**
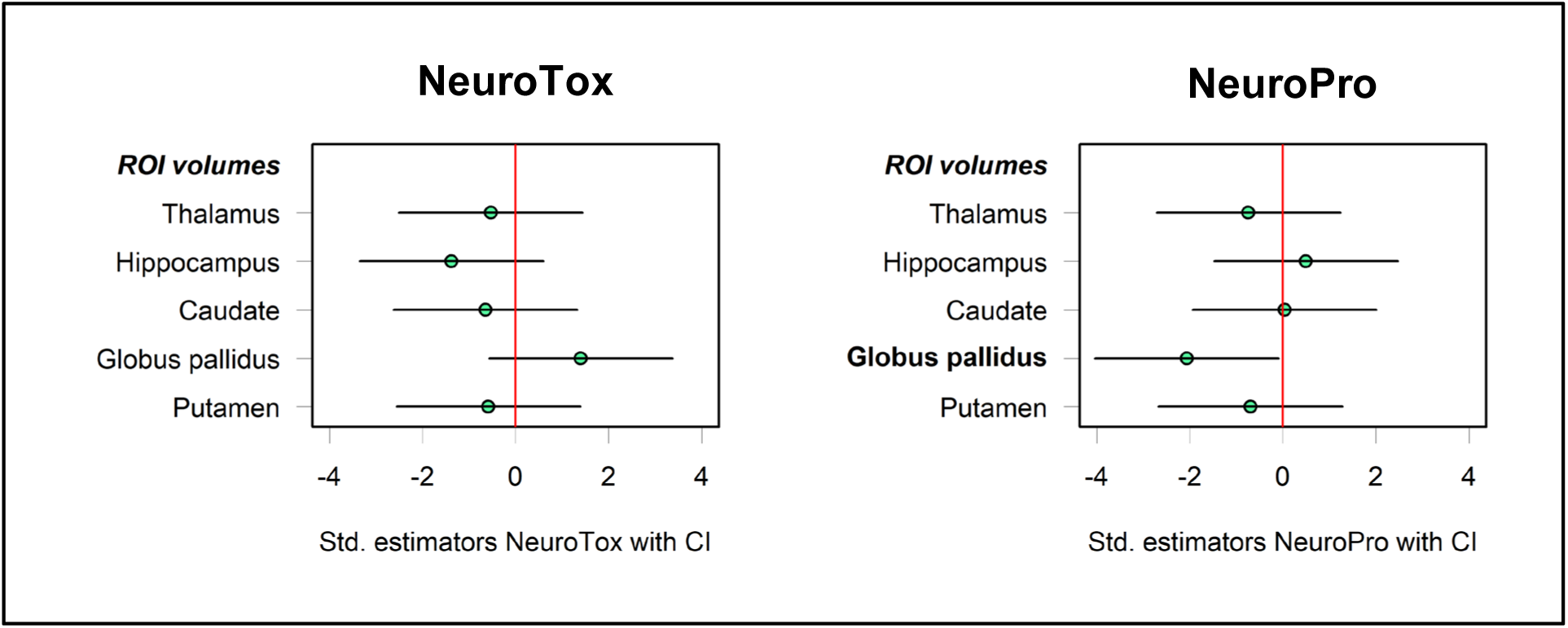
Associations between the kynurenine pathway metabolite patterns “NeuroTox” and “NeuroPro” with regional volumes of subcortical brain structures. Estimator plots showing the standardized estimators of the multiple regression models with “NeuroTox” and “NeuroPro” as predictor variables and MRI metrics (ROI volumes) as predicted variables based on pooled data from the EMSES, EXBRAIN, and PoTOMS studies. ROI = region of interest.

